# Emergence and co-circulation of Monkeypox virus Clade Ia and Clade Ib in South Kivu, Democratic Republic of the Congo, January–May 2026

**DOI:** 10.64898/2026.06.25.26356562

**Authors:** Leandre Murhula Masirika, Jean Claude Udahemuka, David F. Nieuwenhuijse, Bertin Chasinga, Reuben Sindayiheba, Leonard Schuele, Hayley Cassidy, Francisco Bacon Benimana, Akili Chigabo, Jacques Bihando, Gisèle Nzigire Barhatwira, Justin Bengehya Mbiribindi, Jules Ndoli Minega, Trudie Lang, Willy Lulihoshi Kasi, Polepole Ngabo, Stephanie Mitchell, Christian Gortazar, Frank M. Aarestrup, Esto Bahizire, Marion Koopmans, Bas B. Oude Munnink, Pacifique Ndishimye

## Abstract

In September 2023, the first infections with a novel lineage of mpox were detected in South Kivu. Since then, the virus has spread regionally, nationally and internationally. As part of continued efforts to understand the mpox ecology and epidemiology, the South Kivu district of public health and partners have set up systematic case finding and follow-up, including strain characterisation through PCR and sequencing. Samples were collected from 595 hospitalized patients with a confirmed mpox virus infection. A clade differentiating RT-PCR showed that 545 (92%) of samples were positive for clade Ib but also - remarkably- that Clade Ia infections were diagnosed for the first time in South Kivu. First detected in cases in week 7 in Kamituga, 50 cases were identified over the whole study period (8,40% of all cases). Phylogenetic analysis of initial cases revealed introductions of clade Ia into the South Kivu province alongside the continuation of the clade Ib mpox virus outbreak. These findings underscore the increasing complexity of clade I mpox virus outbreaks in the DRC.

## Introduction

Human mpox (formerly monkeypox) is an emerging zoonotic disease caused by mpox virus (MPXV) which is enzootic in Central and West African forested areas [1]. MPXV has become a national reportable disease in the Democratic Republic of Congo (DRC) which is considered as the worldwide epicenter of MPXV [2] and, where the first recorded human case of MPXV was reported in 1970 in a nine-month-old child at Basankusu Hospital [4]. Afterwards cases of MPXV linked to wildlife reservoir exposures followed by local sporadic epidemics have been reported continually in the central and west parts of DRC and in the neighboring countries with limited further geographical spread, however this changed after the global 2022 outbreak of mpox [5,6,7].

Two genetically distinct clades of MPXV have been described with each respectively divided into subclades [8]. Clade I MPXV are historically mainly described in Central and Eastern Africa while clade II MPXV were mainly described in Western Africa [9]. Initially limited research was performed on MPXV but this changed when MPXV gained global attention due to its emergence outside of its endemic region, with a significant outbreak of mpox virus clade IIb in 2022 [10]. This global outbreak spread predominantly – and continues to spread mainly within sexual networks of men who have sex with men [11].

While the clade IIb virus outbreak was resurging, between 2024 and 2025 a sharp increase in MPXV clade I cases was observed throughout all provinces of DRC and mpox virus was detected in areas where it had not been described before, Particularly the South-Kivu province, where the newly reported Clade Ib was detected and heavily affected [12, 13,14,21]. This new clade resulted in a sustained human-to-human epidemic, primarily by (hetero)sexual transmission [14,15], leading to the second declaration of a Public Health Emergency of International Concern (PHEIC) in 2024 by the World health Organization [18]. Recently, the co-occurrence of multiple subclades (Clade Ia, Clade Ib and Clade IIb) were reported in Kinshasa indicating the increasing complexity of mpox outbreaks [16,20]. Co-circulation of different mpox clades also increases the potential for generation of novel recombinant viruses which has already been described once [17]

The geographic range of Clade Ib and Clade IIb is currently widespread [18,19] with cases reported in historically non-endemic countries [20]. The province of South-Kivu presents a particular concern, being the epicenter of the start of the outbreak of the Clade Ib MPXV, and considering its proximity to Rwanda and Burundi, with international connections via the international airports of Kigali and Bujumbura.

Here, we report the first MPXV clade Ia cases followed by rapid spread in the Province of South-Kivu revealing further changing epidemiology and increasing co-circulation of MPXV strains of both subclades, Ia and Ib, in the seven South Kivu health zones Kamituga, Kadutu, Kaziba, Nyangezi, Nyantede, Miti-Murhesa, Kalehe, between January 1 and May 9, 2026.

## Methods

### Case definitions

A case was classified as a “suspected case” if acute symptoms such as fever, severe headache, and muscle and back pain were present, followed by a rash that developed within one to three days, often starting either on the face or genital area before spreading to the rest of the body. A confirmed case of mpox was defined as being diagnosed in the laboratory by an mpox specific real-time PCR. A case was classified as a “probable case” if it met the clinical criteria for a suspected case and had an epidemiological link to a confirmed or probable case but was not confirmed by laboratory tests [27].

### Study population

The study involved patients from South Kivu province in the territories of Mwenga, Walungu, Kabare, Kalehe and Uvira who were hospitalised in the Kamituga, Kamanyola, Nyante nde, Karhanda, Miti, Kalehe, Kaziba, Fomulac Katana, Uvira and Minova hospitals, within Kamituga, Nyangezi, Nyantende, Miti-murhesa, Katana, Kaziba, Ibanda, Kadutu, Bagira, Minova, Kalehe and Uvira health zones (Figure2).

**Figure 1.**
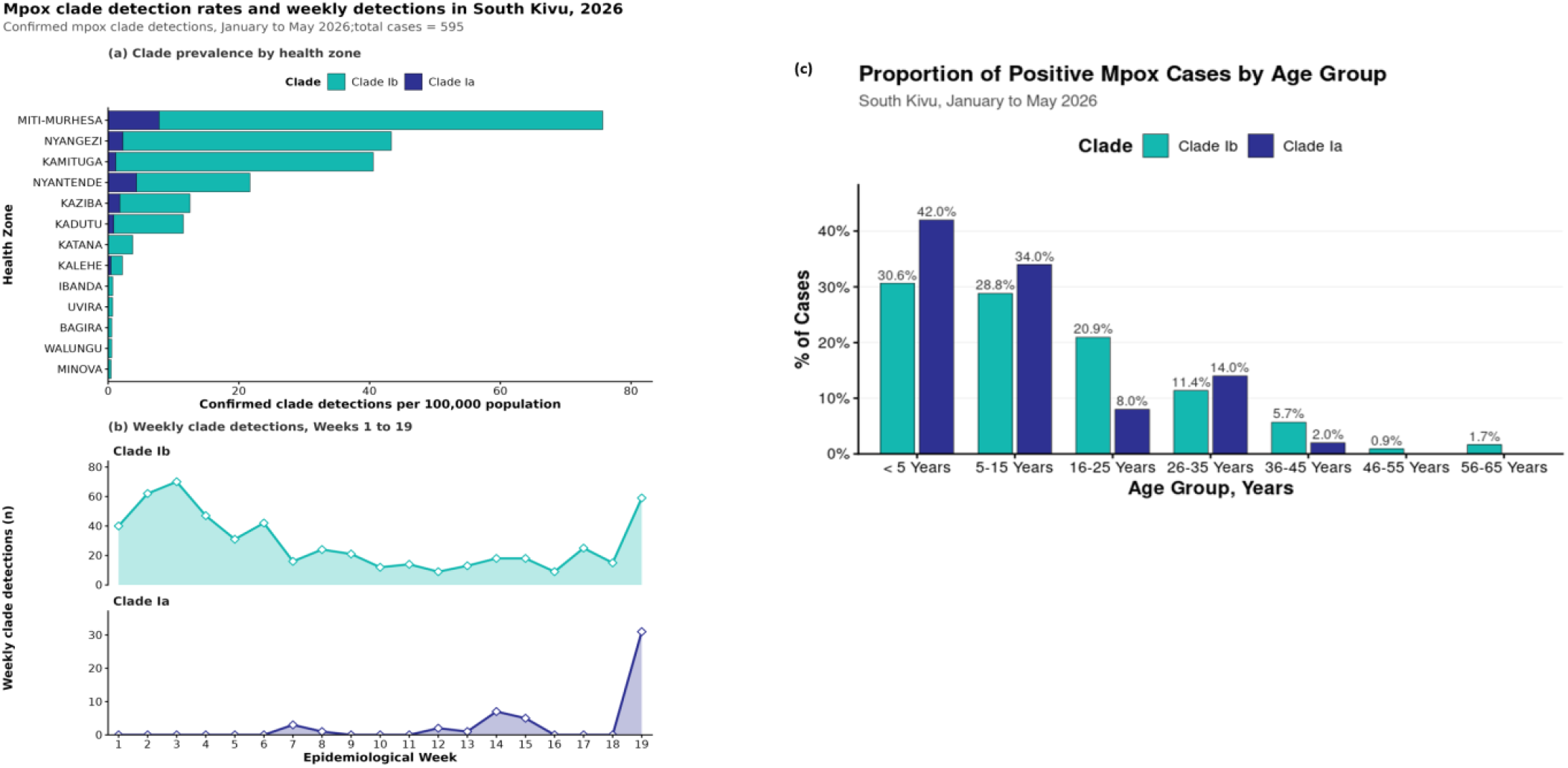
Mpox clade detection rates and weekly clade detections in South Kivu Province, Democratic Republic of the Congo, January to May 2026. Panel A shows cumulative confirmed mpox clade detections per 100,000 population by health zone. Bars are stacked by clade, with Clade Ib shown in teal and Clade Ia shown in blue. Dual-positive Ia/Ib records were counted once under each detected clade. Panel B shows weekly confirmed clade detections from epidemiological weeks 1 to 19. Panel C shows the age distribution of cases. Remarkably, of the 50 confirmed cladeIa, 37(74%) individuals were dual positive for clade Ia and clade Ib and only 13(26%) were positive to clade Ia only as showm the sumplements table3.

**Figure 2.**
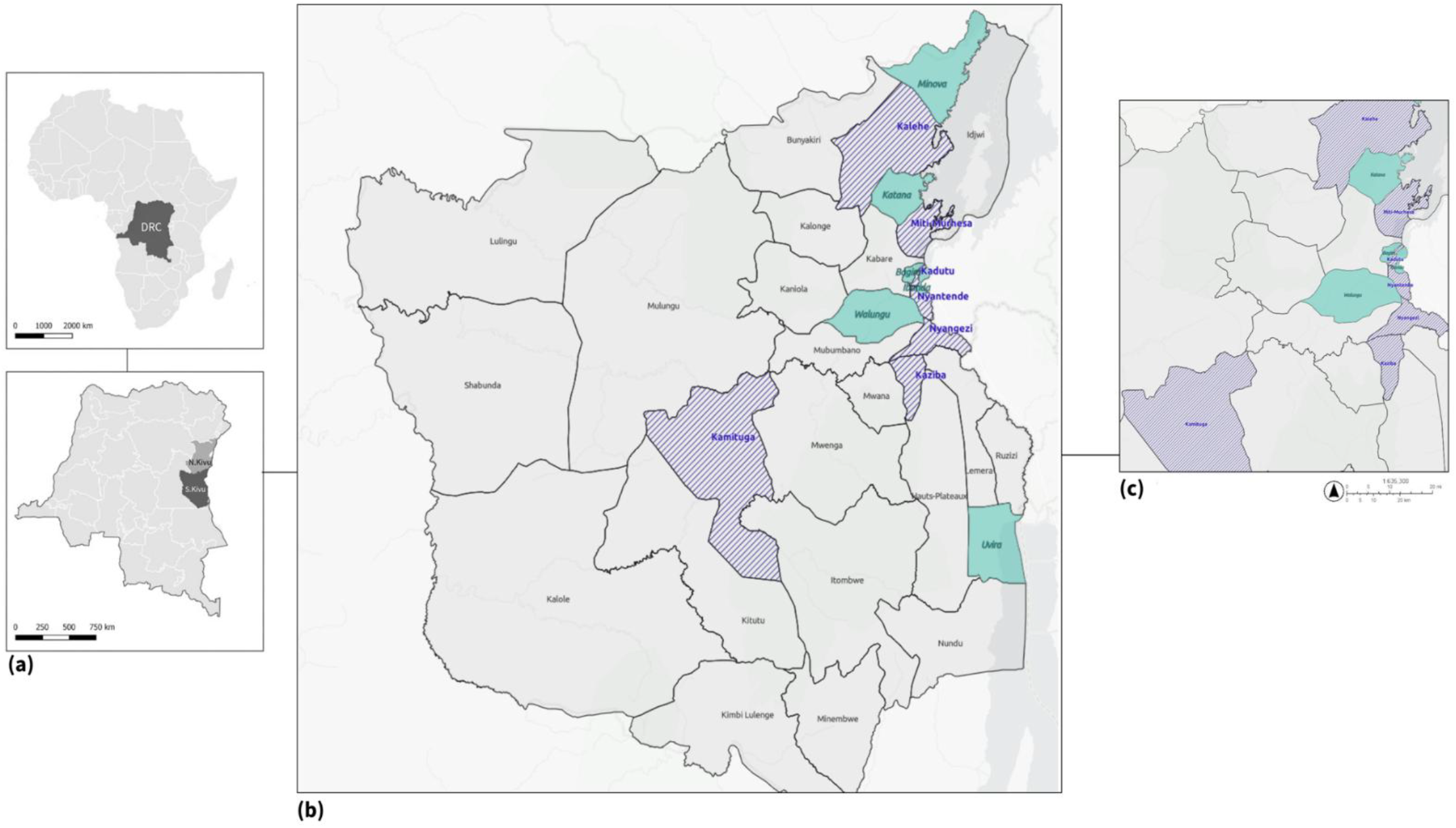
Spatial distribution of confirmed mpox clade detections in South Kivu Province, Democratic Republic of the Congo, 2026. **Panel A** shows the regional context, including the location of the Democratic Republic of the Congo within Africa and South Kivu Province within the DRC. **Panel B** shows the South Kivu outbreak by health zone. **Panel C** provides a closer view of the health zones with the highest number of clade detections. **Green** health zones indicate areas where only Clade Ib was detected. **Blue-striped** health zones indicate areas where both Clade Ia and Clade Ib were detected. *No health zones had Clade Ia detections only*.

### Sample Collection and laboratory procedures

The samples have been collected by local clinicians who provided data from each suspected case using the national investigation form. This form includes information on demographic characteristics (age, sex, place of residence, including health zone and province, occupation and nationality), date of onset of clinical symptoms, type of sample, and date of sample collection. These were recorded into a secured, anonymized database. Admission to the hospitals was based on clinical diagnosis of mpox by hospital staff. According to routines of mpox collection, either vesicles (n = 402), pustules (n = 11), crusts (n = 163) or oropharyngeal swabs (n = 19) were collected from the patients between 1st January to 9th May 2026.

We analyzed routinely collected surveillance and newly generated genomic data from confirmed mpox cases in South Kivu Province. Cases were laboratory-confirmed at the Division Provinciale de la Santé of Sud-Kivu (Laboratory of DPS-South-Kivu) using a clade specific PCR assay (RADIONE Multipox Detection Kit (RP029, KH Medical, Seoul, Republic of Korea) according to the manufacturer’s protocol), and a subset of samples with CT values below 30 and with sufficient sample volume were selected for genomic sequencing. Case confirmation was categorized by clade (Clade Ia and Clade Ib). Cycle values (Ct) were used as semi-quantitative estimates of viral concentrations.

### Genomic sequencing and analysis

The samples from the 595 patients had been collected throughout the month of January– May 2026. Samples were stored in virus transport medium and frozen at −20 °C. For whole-genome as performed on early (first 3 cases) of the cladeIa outbreak collected in Kamitiga health zone. DNA was extracted with the Blood and Tissue Kit (Qiagen) according to the manufacturer’s recommendations. Viral transport medium was used as a negative control. MPXV DNA was amplified using a mpox amplification scheme specifically designed for Clade IIb MPXV to generate 2,500 bp amplicons scheme (https://github.com/quick-lab/primerschemes/tree/main/primerschemes/artic-inrb-mpox/2500/v1.0.0) and quantified using the Qubit dsHS DNA assay (ThermoFisher). Sequencing libraries were prepared using the Native Barcoding Kit 24 V14 (SQK-NBD114.24, Oxford Nanopore Technologies; ONT) and sequenced using a R10.4.1 flow cell.

Sequencing reads were basecalled using Dorado basecalling software version 0.6.2 on high accuracy setting. Raw reads were quality controlled using fastp (version v1.3.3), primer sequences were trimmed using Ampliclip (https://github.com/dnieuw/Ampliclip), and consensus sequences were generated using Virconsens (https://github.com/dnieuw/Virconsens) at a 10x depth threshold. Mutations at less than 30x coverage were manually checked for validity. Consensus sequences were aligned and masked using Squirrel (version 1.3.2) using a manually curated new Clade Ia masking strategy (https://github.com/dnieuw/mpox-genomics-cladeIa-phylomasking). A maximum likelihood phylogenetic tree was generated using IQ-TREE (version 3.1.1) with parameters ‘-m HKY -blmin 0.0000000001 -czb -asr -o ‘KJ642615’ -B 1000’. APOBEC3 mutations were annotated on the branches using a custom python script. Visualisations were made using Peartree v1.2.0 and Inkscape v1.4.

### Mapping

Geolocation and epidemiological data were processed using the R. The epidemiological curve was plotted using ggplot2. Descriptive maps were generated using ArcGIS software. For the geographical maps, cases distribution by population density and aggregated by health area are defined by the shapefiles provided by the national health ministry in the DRC available at https://data.humdata.org/dataset/drc-health-data

### Clade-specific epidemic growth model

The data represent observed outbreak detections over time rather than transmission in a fully susceptible population at the start of the outbreak, therefore, we estimated clade-specific reproduction measures from observed epidemic dynamics: a growth-rate-based reproduction estimate in the primary analysis and an outbreak-average renewal-equation estimate as a sensitivity analysis [26]. Records with both clade Ia and clade Ib detected were counted once under each detected clade, resulting in 595 total clade detections: 545 clade Ib detections and 50 clade Ia detections. We used an exponential growth model to estimate whether clade-specific detections increased or decreased over the observation period and were less sensitive to sparse, late-clustered detections than renewal-equation estimates. For each clade, daily detections were modeled using a generalized linear model with a log link:

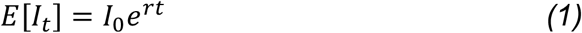

where *I*_*t*_ is the number of clade detections on day *t, I*_0_ is the initial expected detection rate, *α* is the intercept, and *r* is the daily exponential growth rate. Positive values of *r* indicate increasing detections, while negative values indicate declining detections. We converted estimated growth rates to approximate reproduction numbers using a gamma-distributed generation interval with mean *μ* and standard deviation *σ*:

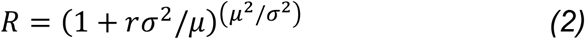

*Doubling time for positive growth was calculated as:*

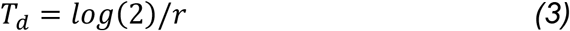

*Halving time for negative growth was calculated as* :

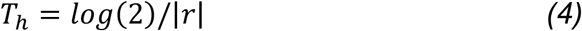

Uncertainty was summarized using 95% confidence intervals for the growth rate and corresponding reproduction number estimates. We also estimated outbreak-average renewal-equation reproduction numbers for each clade for sensitivity (Supplement2) as renewal estimates were more sensitive to sparse clade-specific time series, particularly for Clade Ia.

## Results

### 1. Patients Characteristics by clades

In total, 595 cases were diagnosed. Of these, 545 (91,60%) tested positive for clade Ib and 50 (8,4%) for clade Ia. The clade detections per 100.000 persons per health zone and the cumulative number of cases per clade are shown in Figure 1a and 1b, respectively. Detailed descriptions are given in the supplementary1 tables 1-3. Cases were distributed across 13 health zones (Figure 2). The Miti-murhesa health zone accounted for the majority of cases of both clades, with 226 cases (41,47% of the total number of clade Ib cases) tested positive for clade Ib and 26 cases (52% of the total number of clade Ia cases) tested positive for clade Ia. This was followed by Kamituga health zone with 101 cases of clade Ib (18,53%) and 3 cases of clade Ia (6%) and the Nyanyezi health zone with 91 cases of clade Ib (16,70%) and 5 cases of clade Ia (10%).

Age distribution was bimodal to both clades with more individuals younger than 5 years for both clades, with 21(42%) for clade Ia and 167(30,06%) for clade Ib followed by individuals aged 5-15 Figure 1C. In total, 295 (54,13%) of clade Ib cases were female, and 250 (45,87%) were males. For clade Ia, the distribution was 28 (56 %) females, compared to 22 (44%) males. Regarding profession, we observed a shift in term of transmission compared with our previous reports [14,15]. Here, pupils were the most affected, representing 252 (42,35%) followed by individuals with no profession,185 (31,09%). Traders and farmers represented 58 (9,75), 49 (8,24%) respectively. 33 (5,55%) mining workers and 18 health workers representing (3,03 %), reporting that the co-circulation of cladeIa and cladeIa was heavily linked to close intra-household contacts and school exposures.

### 2. Mapping cases distribution of monkeypox virus’s subclade Ia and Ib

### 3. Clade specific epidemic growth model

The modelling of clade specific growth showed diverging trends for Clade Ia and Ib. In the primary growth-rate model, the estimated daily growth rate for Clade Ia was positive (*r* = 0.0426, 95% CI : 0.0238 to 0.0615), corresponding to an estimated reproduction number > 1 (*R* = 1.44, 95% CI: 1.23 to 1.67). The estimated doubling time was 16.3 days. In contrast, Clade Ib showed a slightly negative daily growth rate (*r* = −0.0067, 95% CI: −0.0113 to −0.0022), corresponding to an estimated reproduction number below 1 (*R* = 0.94, 95% CI: 0.90 to 0.98). The estimated halving time was 102.9 days, indicating a gradual decline in Clade Ib detections over the observation period. Clade Ia detections increased over time. Details are shown in the supplemantary 2.

### 4. Phylogenetic analysis

Near completed genomes were generated for the first 3 cases diagnosed as Clade Ia. This provided evidence for 2 independent sequence clusters. The sequence shown in red in the top part of the phylogenetic tree in Figure 3 is embedded in a larger cluster of sequences related to outbreaks in Tshopo province. The sequences in red in the bottom part of the tree cluster shows a similar picture, although sequences from other provinces are intermixed.

**Figure 3.**
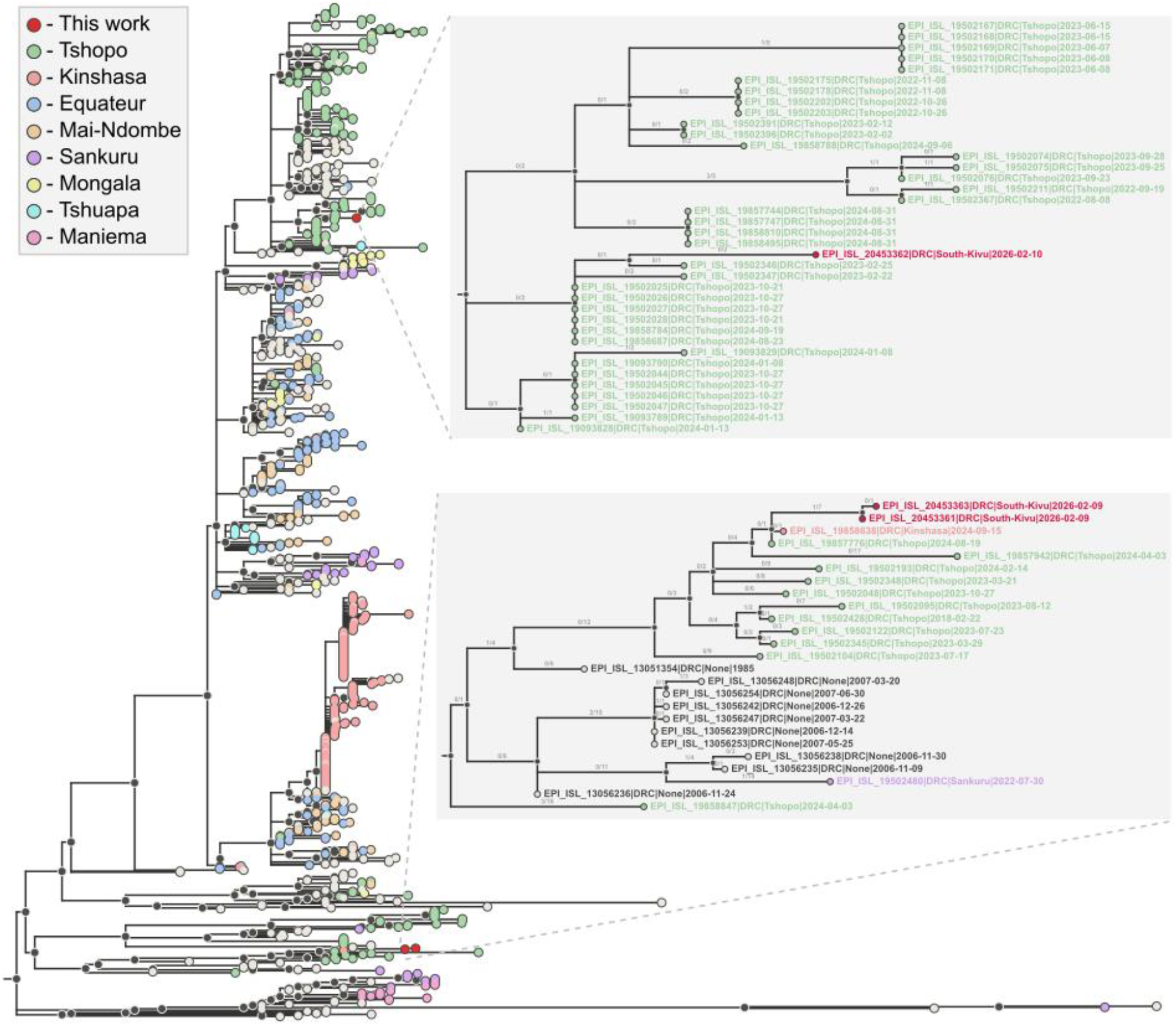
Maximum likelihood phylogenetic tree of all complete mpox Clade Ia sequences available in GISAID. Sequences from the 8 top most abundant provinces are colored, Newly generated near complete genomes are highlighted in red, others are gray/black. Zoom-in of the two clusters are displayed on the right. APOBEC3 mutations are indicated as “APOBEC3/all mutations” on the branches of the zoom-ins.

## Discussion

This report documents the first confirmed mpox Clade Ia cases identified alongside with continued clade Ib cases in South Kivu Province. Prior to 2026 no formal documented case of clade Ia was reported in South-Kivu. We now report the first cases of clade Ia co-circulating alongside with a continued outbreak of clade Ib in 13 health zones. In 2023, the first clade Ib cases were identified in Kamituga health zone primarily associated with (hetero) sexual networks with Profesional Sex Workers (PSW)[14]. Remarkably, these first clade Ia cases in South-kivu are also reported in Kamituga from February 2026 onwards. Clade Ib spread was linked to bars within PSW in densely areas and extended to other health zones through the movements of populations for mining activities)[15]. The current situation is different as most cases reported are (young) children for both clade Ia and clade Ib [29,30]. Clade Ib mpox exhibits mutations driven by human APOBEC-3 enzymes [14,31,32] which suggested a human to human transmission the ongoing clade Ib outbreak. It is unclear if the shift in age range for Clade Ib is a reporting bias or a real shift, in which case it suggests a change in the epidemiology towards more household or community transmission. For the clade Ia cases, the relatively young age of individuals infected has historically been observed for primary zoonotic spillovers. However, clade Ia had previosuly not been observed in South Kivu, despite enhanced surveillance since the Clade Ib epidemic, again suggesting a change in epidemiology.The clade Ia sequences had no evidence of APOBEC mutations, - previosulty reported as evidence for sustained human-to-human transmission, but the epidemiological data suggest that the increasing number of clade Ia cases reflects community exposures (schools). We are currently characterising strains from cases throughout the period to gain a better understanding of the situation.

The eastern region of the DRC has been plagued by a persistent armed conflict since the 1990s, which has severely weakened health systems and individual access to basic healthcare [25]. The co-circulation of clades Ia and Ib, combined with the recent declaration of the Bundibugyo strain of the Ebola virus in South Kivu, complicates outbreak surveillance in the DRC. In addition, armed groups have illegally occupied the Kahuzi-Biega National Park (PNKB), weakening park surveillance and encouraging illegal bushmeat consumption by local communities. In this context, the risk of emergence of new zoonotic spillover may be related to these events. Nevertheless, surveillance of mox outbreaks is still ongoing and we cannot ignore undetected cases in other health zones. More sequences are needed to understand possible human-to-human transmission within the genome of clade Ia, and the implementation of a contact tracing is essential for further investigations.

## Conclusion

This report describes a co-circulation in South-Kivu of two different MPXV subclades, Ia and Ib, between January and May 2026 with a rapid surge in the number of cases of MXPV clade Ia infections in recent weeks.Ongoing genomic studies are expected to provide further information on the circulation of these subclades in different health zones and regions. Intensive surveillance and additional epidemiological research within communities are needed to better understand and control the factors contributing to MPXV outbreaks.

## Data Availability

Newly generated mpox Clade Ia sequences are available in GISAID under accession numbers EPI_ISL_20453362, EPI_ISL_20453363, EPI_ISL_20453361. To access the epidemiological informations related to the manuscript please contact

## Acknowledgements

We would like to thank the Provincial Division of Health (DPS) of South-Kivu and all Health Zones from South Kivu for their collaboration during the study. We greatly thank Wildlife Conservation Network (WCN) for the scholarship and research support they awarded to the first author. We acknowledge the UK Medical Research Council (MRC) and the UK Foreign, Commonwealth & Development Office (FCDO) for their research support under the MRC-UKRI grant MC_PC_24001, which was awarded to local investigators, Kamituga Hospital, and the Division Provinciale de la Santé (DPS, Provincial Division of Health) in South Kivu, Bukavu. Finally, the DPS acknowledges the support of the Bill & Melinda Gates Foundation [INV-078955 AIMS-NEI] for grant awarded to local clinians investigators, in South-Kivu Hospitals, for the research on mpox and pregancy outcomes. The funders had no role in study design, data collection and analysis, decision to publish, or preparation of the manuscript.

## Ethical clearance

The ethical clearance to conduct this study was obtained from the Ethical Review Committee of the Catholic University of Bukavu (Reference Number : UCB/CIES/PB/GM/015/2025). All study participants provided informed consent or in the case that the participant was a minor, parental permission or assent was obtained.

## Authors’ contributions

All authors approved the final version of the paper. LMM, JCU, PN, BBOM, and MK. conceptualized and designed the study. MM, JCU, BBOM, PN, FMA, TL, DFN, SM,RS,FBB, GZB,AC,JB,JBM,KLW,PN and MK contributed to data acquisition, LMM,HC,LS,EB,BC,JNM,SM,BBOM, DFN,CG,PN,JCU and MK drafted the paper and figures. LMM,JBM,and KWL collected epidemiological data and arranged for patient sampling. All authors were involved in paper writing and approved the final version.

## Data availability

**Newly generated mpox Clade Ia sequences are available in GISAID under accession numbers EPI_ISL_20453362, EPI_ISL_20453363, EPI_ISL_20453361**.

