## Supplementary Information1 for "Emergence and co-circulation of Monkeypox virus Clade Ia and Clade Ib in South Kivu, Democratic Republic of the Congo, January–May 2026"

### Renewal-equation sensitivity analysis

As a sensitivity analysis, we estimated an outbreak-average reproduction number using a renewal-equation framework. This analysis was conducted separately for Clade Ib and Clade Ia using daily confirmed mpox clade detections from South Kivu Province, Democratic Re-public of the Congo, from January to May 2026. Records with both Clade Ia and Clade Ib detected were counted once under each detected clade. For each clade, expected infectiousness at time *t* was calculated from previous detections weighted by the assumed generation interval distribution:

$$\lambda_t = \sum_{s=1}^{t-1} I_{t-s} w_s$$

where $\lambda_t$ is total infectiousness at time $t$, $I_{t-s}$ is the number of detections $s$ days earlier, and $w_s$ is the generation interval probability at lag $s$. We then estimated an outbreak-average reproduction number as the ratio of observed detections to total expected infectiousness:

$$R = \frac{\sum_t I_t}{\sum_t \lambda_t}$$

We interpreted these renewal-equation estimates as sensitivity analyses rather than primary estimates because the method assumes that observed detections arise from prior clade-specific infectiousness. This assumption may be less appropriate for sparse clade-specific detection series, particularly if some detections reflect independent introductions rather than a single sustained transmission chain. Because Clade Ia detections were relatively few and concentrated later in the observation period, renewal-equation estimates for Clade Ia were interpreted cautiously. The renewal-equation sensitivity analysis produced higher outbreak-average reproduction number estimates than the primary growth-rate model. Clade Ib had an outbreak-average renewal estimate of $R$ = 1.11, with an uncertainty interval of 1.02 to 1.21. Clade Ia had an outbreak-average renewal estimate of $R$ = 2.33, with an uncertainty interval of 1.73 to 3.02. These estimates were directionally consistent with ongoing clade-specific transmission but differed from the primary growth-rate estimates, particularly for Clade Ia. The higher Clade Ia estimate likely reflects the sensitivity of renewal-equation approaches to sparse detections occurring after periods with little prior clade-specific infectiousness.
