## Supplementary Information2 for "Emergence and co-circulation of Monkeypox virus Clade Ia and Clade Ib in South Kivu, Democratic Republic of the Congo, January–May 2026"

**Table 1. numbers of cases for cladeIa and cladeIb within health zones South Kivu, 2026, January to May 2026, Weeks 1 to 19**

| Health Zones | CladeIa | CladeIb | CladeIa % | CladeIb % |
|---|---|---|---|---|
| BAGIRA | 0 | 1 | 0,00% | 0,18% |
| IBANDA | 0 | 4 | 0,00% | 0,73% |
| KADUTU | 4 | 50 | 8,00% | 9,17% |
| KALEHE | 1 | 4 | 2,00% | 0,73% |
| KAMITUGA | 3 | 101 | 6,00% | 18,53% |
| KATANA | 0 | 11 | 0,00% | 2,02% |
| KAZIBA | 3 | 18 | 6,00% | 3,30% |
| MINOVA | 0 | 2 | 0,00% | 0,37% |
| MITI-MURHESA | 26 | 226 | 52,00% | 41,47% |
| NYANGEZI | 5 | 91 | 10,00% | 16,70% |

| | | | | |
|---|---|---|---|---|
| NYANTENDE | 8 | 32 | 16,00% | 5,87% |
| UVIRA | 0 | 3 | 0,00% | 0,55% |
| WALUNGU | 0 | 2 | 0,00% | 0,37% |
| **Total** | **50** | **545** | **100%** | **100%** |

**Table 2. Weekly mpox clade detections, South Kivu, 2026, January to May 2026, Weeks 1 to 19**

| ***Epiweek*** | **Weekly (Ib)** | **Weekly (Ia)** | **Total (Ib)** | **Total (Ia)** | **Zones (Ib)** | **Zones (Ia)** |
|---|---|---|---|---|---|---|
| ***1*** | 40 | 0 | 40 | 0 | KADUTU, KAMITUGA, MITI-MURHESA, NYANGEZI, NYANTENDE | None |
| ***2*** | 62 | 0 | 102 | 0 | KADUTU, KAMITUGA, MITI-MURHESA, NYANGEZI, NYANTENDE | None |
| ***3*** | 70 | 0 | 172 | 0 | KADUTU, KAMITUGA, MITI-MURHESA, NYANGEZI | None |
| ***4*** | 47 | 0 | 219 | 0 | KADUTU, KALEHE, KAMITUGA, KAZIBA, MITI-MURHESA, NYANGEZI, NYANTENDE | None |
| ***5*** | 31 | 0 | 250 | 0 | KADUTU, KALEHE, KAMITUGA, KAZIBA, MITI-MURHESA, NYANGEZI, NYANTENDE | None |
| ***6*** | 42 | 0 | 292 | 0 | BAGIRA, KADUTU, KAMITUGA, MITI-MURHESA, NYANGEZI, NYANTENDE | None |
| ***7*** | 16 | 3 | 308 | 3 | KADUTU, KAMITUGA, MITI-MURHESA, NYANGEZI | KAMITUGA |
| ***8*** | 24 | 1 | 332 | 4 | KALEHE, KAMITUGA, KATANA, MITI-MURHESA, NYANGEZI | KALEHE |
| ***9*** | 21 | 0 | 353 | 4 | KADUTU, KAMITUGA, KATANA, MINOVA, MITI-MURHESA, NYANGEZI | None |

| | | | | | | |
|---|---|---|---|---|---|---|
| ***10*** | 12 | 0 | 365 | 4 | IBANDA, KADUTU, KAMITUGA, MITI-MURHESA | None |
| ***11*** | 14 | 0 | 379 | 4 | KAMITUGA, KATANA, MITI-MURHESA, NYANGEZI | None |
| ***12*** | 9 | 2 | 388 | 6 | KAMITUGA, MITI-MURHESA | MITI-MURHESA |
| ***13*** | 13 | 1 | 401 | 7 | IBANDA, KAMITUGA, MITI-MURHESA, NYANGEZI | MITI-MURHESA |
| ***14*** | 18 | 7 | 419 | 14 | KAMITUGA, KAZIBA, MITI-MURHESA, NYANGEZI | MITI-MURHESA, NYANGEZI, NYANTENDE |
| ***15*** | 18 | 5 | 437 | 19 | KAMITUGA, MITI-MURHESA, NYANGEZI, UVIRA | MITI-MURHESA |
| ***16*** | 9 | 0 | 446 | 19 | KAMITUGA, MITI-MURHESA, NYANGEZI | None |
| ***17*** | 25 | 0 | 471 | 19 | KAMITUGA, MITI-MURHESA, NYANGEZI, WALUNGU | None |
| ***18*** | 15 | 0 | 486 | 19 | IBANDA, KAMITUGA, MITI-MURHESA, WALUNGU | None |
| ***19*** | 59 | 31 | 545 | 50 | KADUTU, KAMITUGA, KAZIBA, MITI-MURHESA, NYANGEZI, NYANTENDE | KADUTU, KAZIBA, MITI-MURHESA, NYANGEZI, NYANTENDE |

**Table 3: Demographics 9and weekly outbreak summary, South Kivu, 2026, January to May 2026, Weeks 1 to 19**

| *Week* | Total clade detections | Ib detections | Ia detections | Ib only | Ia only | Dual-positive | Median age | Age IQR | % <15 yrs |
|---|---|---|---|---|---|---|---|---|---|
| *1* | 40 | 40 | 0 | 40 | 0 | 0 | 13.0 | 3.8 to 22.2 | 55.0 |
| *2* | 62 | 62 | 0 | 62 | 0 | 0 | 6.0 | 3 to 20 | 64.5 |
| *3* | 70 | 70 | 0 | 70 | 0 | 0 | 7.0 | 2.2 to 24.8 | 67.1 |
| *4* | 47 | 47 | 0 | 47 | 0 | 0 | 13.0 | 4.5 to 19 | 55.3 |
| *5* | 31 | 31 | 0 | 31 | 0 | 0 | 13.0 | 6 to 22.5 | 58.1 |
| *6* | 42 | 42 | 0 | 42 | 0 | 0 | 8.5 | 2 to 19 | 57.1 |
| *7* | 19 | 16 | 3 | 13 | 0 | 3 | 14.5 | 6.8 to 22.2 | 50.0 |
| *8* | 25 | 24 | 1 | 23 | 0 | 1 | 11.0 | 4.8 to 27.2 | 58.3 |
| *9* | 21 | 21 | 0 | 21 | 0 | 0 | 20.0 | 8 to 22 | 38.1 |
| *10* | 12 | 12 | 0 | 12 | 0 | 0 | 25.0 | 10 to 40 | 33.3 |
| *11* | 14 | 14 | 0 | 14 | 0 | 0 | 5.0 | 0.8 to 15.8 | 71.4 |
| *12* | 11 | 9 | 2 | 7 | 0 | 2 | 20.0 | 11 to 25 | 33.3 |
| *13* | 14 | 13 | 1 | 12 | 0 | 1 | 23.0 | 15 to 27 | 23.1 |
| *14* | 25 | 18 | 7 | 12 | 1 | 6 | 21.0 | 2.5 to 27 | 42.1 |
| *15* | 23 | 18 | 5 | 13 | 0 | 5 | 15.0 | 9.2 to 29.2 | 50.0 |
| *16* | 9 | 9 | 0 | 9 | 0 | 0 | 20.0 | 2 to 25 | 44.4 |
| *17* | 25 | 25 | 0 | 25 | 0 | 0 | 6.0 | 1 to 16 | 68.0 |
| *18* | 15 | 15 | 0 | 15 | 0 | 0 | 15.0 | 5.5 to 21.5 | 46.7 |
| *19* | 90 | 59 | 31 | 40 | 12 | 19 | 7.0 | 2 to 17.5 | 67.6 |
